# Rapid vaccination and early reactive partial lockdown will minimize deaths from emerging highly contagious SARS-CoV-2 variants

**DOI:** 10.1101/2021.02.02.21250985

**Authors:** Daniel B Reeves, Chloe Bracis, David A. Swan, Mia Moore, Dobromir Dimitrov, Joshua T. Schiffer

**Author notes:** equal contribution (Daniel B Reeves, Chloe Bracis, Dobromir Dimitrov, Joshua T. Schiffer).

## Abstract

The goals of SARS-CoV-2 vaccination programs are to maximally reduce cases and deaths, and to limit the amount of time required under lockdown. Using a mathematical model calibrated to data from King County Washington but generalizable across states, we simulated multiple scenarios with different vaccine efficacy profiles, vaccination rates, and case thresholds for triggering and relaxing partial lockdowns. We assumed that a contagious variant is currently present at low levels. In all scenarios, it rapidly becomes dominant by early summer. Low case thresholds for triggering partial lockdowns during current and future waves of infection strongly predict lower total numbers of COVID-19 infections, hospitalizations and deaths in 2021. However, in regions with relatively higher current seroprevalence, there is a predicted delay in onset of a subsequent surge in new variant infections. For all vaccine efficacy profiles, increasing vaccination rate lowers the total number of infections and deaths, as well as the total number of days under partial lockdown. Due to variable current estimates of emerging variant infectiousness, vaccine efficacy against these variants, vaccine refusal, and future adherence to masking and physical distancing, we project considerable uncertainty regarding the timing and intensity of subsequent waves of infection. Nevertheless, under all plausible scenarios, rapid vaccination and early implementation of partial lockdown are the two most critical variables to save the greatest number of lives.

## Introduction

The COVID-19 pandemic is a profound tragedy that has resulted in widespread death and high morbidity.^1^ Its social impact has also been devastating with recurrent lockdown resulting in economic loss and psychological damage, particularly among children who are being deprived of in-person schooling.^2-4^

Accordingly, recently initiated COVID-19 vaccination programs have public health and societal goals. Vaccine allocation strategies are intended to prevent the largest numbers of cumulative deaths, as well as high peaks in hospitalizations to avoid collapse of healthcare systems.^5^ Because a moderate proportion of infected people develops debilitating long-term symptoms, limiting cumulative case numbers is also a priority.^6^

Another goal is to support conditions allowing sustained safe reopening of schools, businesses, and public events.^7^ One metric for assessing this is the number of days during which partial lockdown will be necessary during 2021. Prior to vaccination implementation, numerous strategies existed to prevent surges in cases and subsequent lockdowns including testing and tracing,^8^ masking,^9,10^ and limiting crowd size in indoor spaces.^11^ While these efforts undoubtedly had positive effects, they were not sufficient to prevent three recurrent waves of infection in the United States.^12^ The first two waves occurred in early spring and mid-summer with the intensity of cases varying substantially among states. The ongoing third wave is more generalized leading to overburdened hospitals and a huge surge in deaths in multiple states.^13^

A further concern is the emergence of more infectious viral variants across the globe with relevant examples in England,^14^ Brazil,^15^ and South Africa.^16^ The B.1.1.7 variant is estimated to be considerably more contagious and is overwhelming many health care systems in the United Kingdom.^17^ B.1.1.7 has also been identified in numerous locations across the United States.

While its prevalence is currently low, B.1.1.7 and potentially other new variants are predicted to predominate in upcoming months.^18^

The precise impact of new SARS-CoV-2 variants in the face of improving non-pharmaceutical preventative measures has yet to be determined. More widespread use of highly efficacious N95 masks, particularly among people who work in potential super-spreader environments, could limit SARS-CoV-2 spread more effectively than current masking approaches.^19^ Widespread availability of inexpensive and rapid tests, paired with effective contact tracing, could also limit incident infections.^20^ However, it remains uncertain whether these additions will be sufficient to prevent exponential growth of the B.1.1.7 variant which increases the effective reproductive number by 56% (range: 50-74%).^21^

During 2020, physical distancing mandates, or partial lockdowns, were often necessary to curb unchecked local epidemics.^22^ The effectiveness of partial lockdowns was evident in the northeast United States and Europe during the spring of 2020, and more recently in multiple northern plains states where incident cases contracted after a very high peak.^13^ In the UK and Ireland, the B.1.1.7 variant emerged during lockdown necessitating a stricter lockdown in January, after which cases have declined.^23^ Unfortunately, partial lockdown measures come at great societal cost, including business and school closures that disproportionately impact socioeconomically disadvantaged regions and communities of color. A major goal of any vaccination strategy should be to limit the duration and intensity of partial lockdown in 2021.

Existing mathematical models have considered optimal allocation of SARS-CoV-2 vaccines,^24-28^ one versus two-dose strategies,^29^ and the impact of vaccine effects against infectiousness.^30-32^ Here, using a model calibrated to data from King County Washington,^33^ we consider the complexities of vaccine implementation during the ongoing intense third wave of SARS-CoV-2 infections, as well as the likely future emergence of more contagious variants. Our goal is to understand which variables have the greatest effect on infections and deaths, in order to limit them while also decreasing total time under lockdown. The model also allows for projections in other states that had lower or higher cumulative incidence at the time when their vaccination programs initiated.

## Results

### King County mathematical model

We developed an epidemiological model of SARS-CoV-2 spread in King County Washington **(Sup fig 1a)**.^30,31,33^ As described elsewhere, we calibrated the model to diagnosed cases, hospitalizations, and deaths, all stratified according to age cohort, to capture the epidemic pre-vaccination through early November.^31^ A mathematical description of the model is presented in the **Methods** and complete mathematical details are found in the **Supplementary Information**.

The model follows three sequential waves of infections with peaks in March, August, and December of 2020. Each wave has different peak incidence and age-cohort predominance and occurred due to an antecedent increase in the effective reproductive number (R_e_).^19^ Without a vaccine, R_e_ is governed by the frequency of exposure contacts between susceptible and infected individuals. A key feature of our model is a time-varying, age-stratified vector (*σ*t) governing social distancing (non-pharmaceutical interventions) including reduced contacts through personal choices and/or mandated partial lockdowns, as well as reductions in exposure contacts due to mask wearing or physical distancing. The components of the vector *σ*t vary from 0, indicating pre-pandemic levels of societal interactivity and no masking, to 1, indicating complete lockdown with no interactions **(Supp Fig 1b)**. For data prior to November 2020, we estimated *σ*t for each age group by calibrating monthly to cases, hospitalization and deaths.

In 2021, we project that *σ*t will continue to vary according to government policy and public behavior. We include several parameters to reflect this uncertainty. The first is the case threshold to trigger partial lockdown (*C*_*max*_). If two-week number of diagnosed cases per 100,000 people rises above *C*_*max*_, a “partial-lockdown” (*σ*_*t*_ → *σ*_*pL*_)is mandated **(Supp Fig 1b)**.

Partial lockdown is defined by an enforced social distancing of *σ* _*pL*_ =0.6 in the 3 non-elderly age cohorts and *σ*_*pL*_=0.8 in the elderly cohort (≥70 years). Values of *σ* _*pL*_ and *C*_*max*_ were selected based on previous partial lockdowns implemented in Washington State during 2020.^31^ We explore a wide range for *C*_*max*_ in our analysis between 200 and 650, both to reflect heterogeneity in severity of the ongoing third wave across states which occurred due to implementing partial lockdowns at varying thresholds, and to represent future uncertainty.

Similarly, *C*_*min*_ is the case threshold (two-week number of diagnosed cases per 100,000 people) which triggers reopening during which *σ*_*t*_ gradually decreases (at 10% every two weeks) from *σ*_*pL*_ to *σ* _*min*_ in the 3 younger cohorts (*σ* _*min*_+0.2 in seniors). We test 20, 60 and 100 as possible values for *C*_*min*_. Unless otherwise noted, the level of social distancing after a period of societal reopening (*σ*_*min*_) is maintained at 0.2 to capture persistent behaviors such as masking, working from home and avoidance of large social gatherings, which inherently limit the number of interpersonal contacts relative to pre-pandemic levels.

### Vaccination scenarios

Three variables measure vaccine efficacy, including protection against infection (VE_SYMP_), against symptoms given infection despite vaccination (VE_SUSC_), and against secondary transmission upon breakthrough infection (VE_INF_).^34^ Each of these vaccine efficacies theoretically ranges from 0-100% (0-1 in our model). Vaccine efficacy against symptomatic disease (VE_DIS)_, which represents a combination of VE_SUSC_ and VE_SYMP_, is usually measured in clinical trials and was estimated at 0.95 for the Pfizer and Moderna licensed vaccines, with unknown contribution by VE_SUSC_ or VE_SYMP_.^35,36^ VE_INF_ could lower secondary transmission considerably if VE_SUSC_ is low, but its values has also not been measured.^31^ Moreover, the efficacy of the Pfizer and Moderna vaccines against new variants remain unknown. The efficacy of the Novavax and Johnson and Johnson products were 90% and 69% against the consensus variant but were less effective against the South Africa B.1351 lineage.^37,38^ We therefore consider low (0.1), medium (0.5) and high (0.9) values for each of the vaccine efficacies.

A final variable is the vaccination rate, *r*. We use a range of possible rates intended to capture national goals (∼3400 people vaccinated per day^7^) and more aggressive state level goals (∼8000 people vaccinated per day^39^) extrapolated to King County. Under all scenarios, we assume current local policy prioritizing seniors by allocating 80% of initial vaccines doses to the elderly cohort until the maximum coverage of 80% among this cohort is reached **(Supp Fig 1c)**.

### Sensitivity analysis

We performed a global sensitivity analysis under a full factorial design of six input variables **(Table 1)**, including two of the described parameters governing social distancing (*C*_*max*_, *C*_*min*_), the three parameters describing vaccine efficacy (*VE*_*SUSC*_, *VE*_*SYMP*_, and *VE*_*INF*_), and vaccination rollout rate *r*, resulting in 3888 vaccination implementation scenarios. We sought to determine which variables carried the most significant impact on COVID-19 related outcomes during 2021 including peak and cumulative infections, diagnosed cases, hospitalization and deaths, as well as total days under partial lockdown (defined by *σ*_*t*_ =0.6 in 3 younger cohorts and *σ*_*t*_ =0.8 in the elderly) and average value for *σ*_*t*_ during 2021. We calculated Spearman’s rank-order correlation, a nonparametric measure assuming monotonicity but not linearity, among outcomes as well as between outcomes and variables.

**Table 1.**
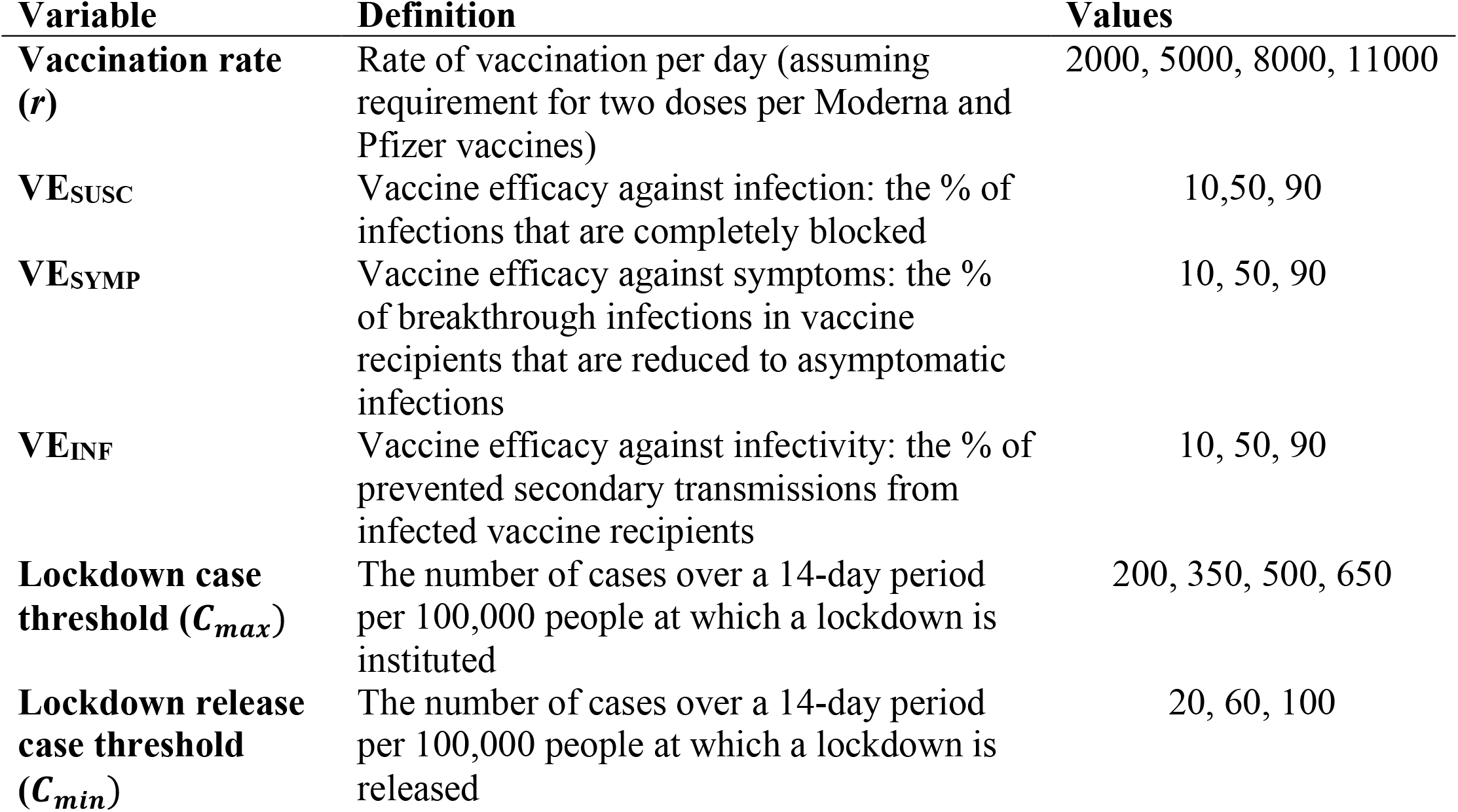
Sensitivity analysis parameters.

### Viral variant assumptions

The SARS-CoV-2 variant B.1.1.7 is widespread in the United Kingdom and has an estimated increase in infectivity of 56% relative to current predominant circulating variants in the US.^21^ On January 23, 2021, the first two cases of the B.1.1.7 variant were detected in nearby Snohomish County WA and are estimated to be ∼1/500 of sequenced samples from a random selection of statewide viruses (Pavitra Roychoudhury, personal communication).^40^ The B.1.1.7 variant is present in multiple other states and widespread cryptic transmission is highly likely. In all simulations presented in the manuscript, we assume two variants, the current circulating consensus and B.1.1.7, which we introduce into the state at a constant rate of five per day beginning January 1, 2021. We also explore scenarios which include plausible bounds of enhanced infectivity (35% and 75% increase), different rates of introduction of the variant into King County (0, 2.5, 5 and 10 per day) and different minimum values social distancing *σ*_*min*_ (0.1 and 0.3).

### High correlation between numbers of infections, hospitalizations and deaths among vaccination scenarios

Various health-related outcomes may be used to assess the success of vaccine distribution including numbers of infected people, diagnosed cases, hospitalizations, and deaths. For each outcome, cumulative totals, totals since vaccination initiation, and peak daily numbers are relevant. Across all simulated scenarios, all health-related outcomes were strongly positively correlated (**Sup fig 2a, 3**). It is therefore unlikely that certain health benefits will be accrued at the expense of others.

### Slight negative correlation between health-related outcomes and total days under partial lockdown among vaccination scenarios

We next considered outcomes that would have societal benefit. Total number of days under partial lockdown correlated negatively with infections and deaths, signifying that vaccine implementation scenarios which most effectively prevent infections and deaths may require longer durations under partial lockdown **(Sup fig 2a, 3)**. The average value of social distancing ⟨*σ σ* ⟩ during vaccine rollout did not correlate with health-related outcomes **(Sup fig 2a, 3)**. This result indicates that complex locally specific epidemiological dynamics govern the relationship between vaccination and social distancing requirements.

### Reduction in COVID-19 infections and deaths due to a low case threshold for triggering partial lockdown

The case threshold for triggering partial lockdown (*C*_*max*_) correlated with all outcomes related to infections, hospitalizations and deaths **(Fig 1a**): assuming a vaccine with high efficacy against infection (VE_SUSC_=90%, VE_SYMP_=10%, VE_INF_=10%) and with national target vaccination rates (3500 per day), a low case threshold for triggering partial lockdown (*C*_*max*_ 200) would substantially limit number of diagnosed cases, infections, hospitalizations and deaths associated with the current third wave **(Fig 1b, blue line)**.

**Figure 1.**
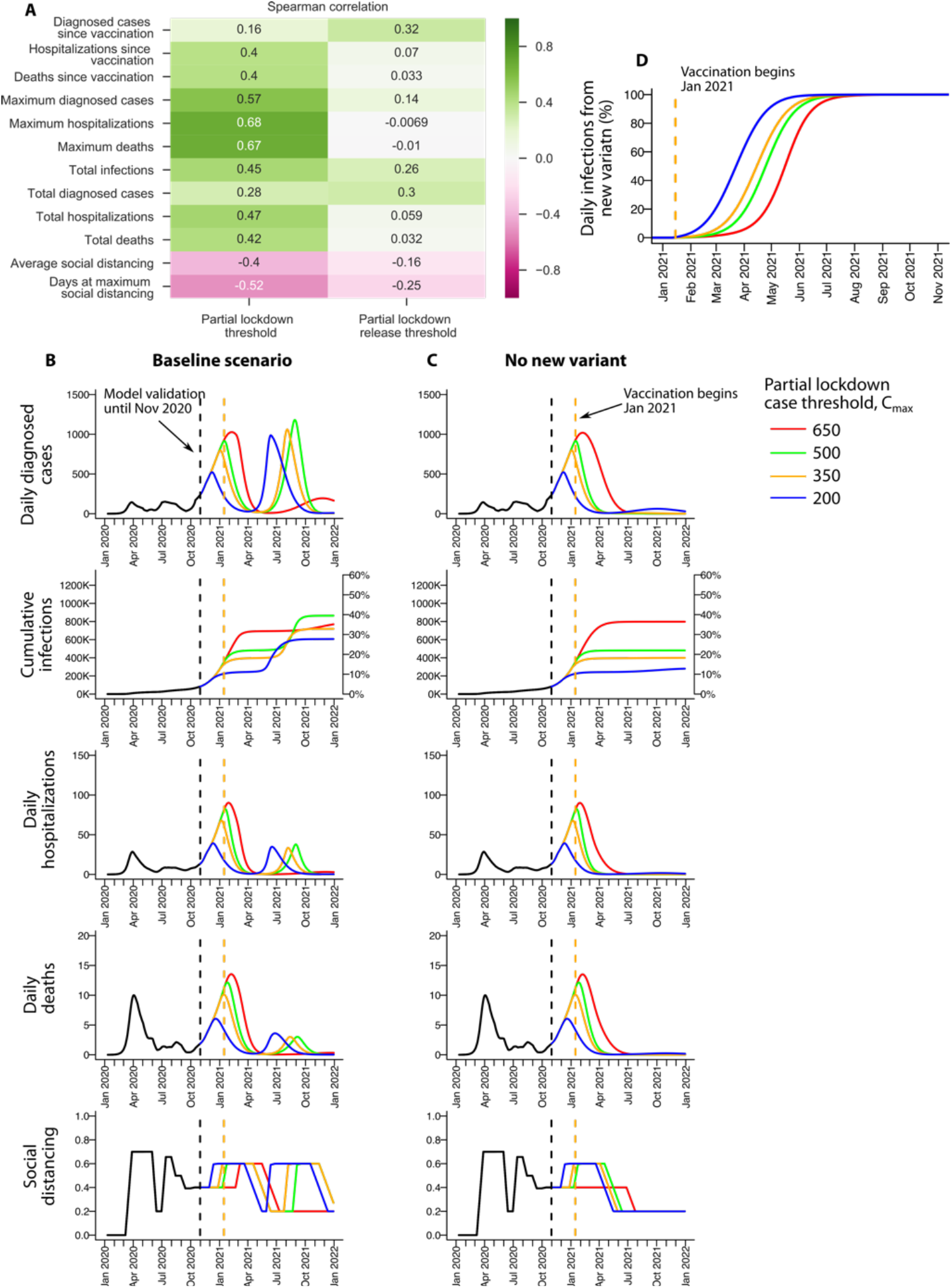
Lowering case threshold for partial lockdown minimizes infections, hospitalizations and deaths but increases total numbers of days under partial lockdown. **a**. Spearman correlations for input variables including case thresholds for triggering (*C*_*max*_) and releasing (*C*_*min*_) partial lockdown. Lowering *C*_*max*_ reduces infections, hospitalizations and deaths, but increases total days at maximal social distancing. *C*_*min*_ has much weaker effects on outcomes. **b**. Impact of varying case threshold for triggering lockdown on relevant outcomes. Vaccination rate is assumed to be 3500 per day with VE_SUSC_=90% / VE_SYMP_=10% / VE_INF_ =10%, *C*_*min*_ =25 cases per 100,000 (2-week average) and *σ*_*min*_ *=*0.2. New variants are present as of January 1 and introduced into the county at a rate of 5 per day. **c**. Equivalent results assuming no new variant. Black dashed vertical line is the end of model calibration. Orange dashed vertical line is the day of vaccine initiation. **d**. Proportion of infections in **b**. due to new variants.

However, due to smaller numbers of infections accumulated during the third wave resulting in a larger proportion of people remaining susceptible to infection, low *C*_*max*_ is projected to be associated with more rapid emergence of a fourth epidemic wave in May 2021, necessitating further partial lockdowns **(Fig 1b, blue line)**. Cumulative infections and deaths are projected to reach 584,311 (27%) and 1524 (0.07%) by 2022. Peak hospitalizations and deaths are expected to be lower during the fourth wave due to early vaccination of the elderly **(Fig 1b)**.

The scenario associated with *C*_*max*_ =350 **(Fig 1b, orange line)** closely reflects current conditions in King County with similar magnitude and timing of peak diagnosed cases, hospitalizations and deaths during the third wave.^41^ Under this assumption, a fourth wave of B.1.1.7 cases is projected in July with a partial lockdown required to stem a surge in infections. Total infections and deaths are projected at 689,057 (31%) and 1806 (0.08%) through 2021.

At high *C*_*max*_ =650, a prolonged, severe third wave is projected through April 2021 leading to 743,578 (34%) of the population infected and 2077 (0.09%) cumulative deaths by 2022 **(Fig 1b, red line)**. Peak per capita diagnosed infections are comparable to third waves in northern plains states that instituted slower lockdowns during recent months.^13^ Because vaccination occurs in the context of fewer susceptible people, partial herd immunity is projected and a delayed and blunted fourth wave is predicted, despite the predominance of the B.1.1.7 variant **(Fig 1b)**.

When we remove “no-lockdown” scenarios with *C*_*max*_ =650 cases per day and thereby assume reactive partial lockdown to high prevalence of local cases, the negative correlation between health-related and societal outcomes across all scenarios disappears. Therefore, reduction in infections and deaths can be prioritized without leading to more days under partial lockdown **(Sup fig 2b)**.

We finally measure the time difference between *C*_*max*_ =200 and 350, 350 and 500, and 500 and 650, to be approximately 2 weeks. Therefore, decisions to institute partial lockdown that are delayed even slightly may substantially increase infections and deaths.

### More rapid predominance of new contagious variants in low prevalence regions

Under all *C*_*max*_ scenarios, the B.1.1.7 variant is projected to account for a majority of cases during the fourth wave of cases **(Fig 1d)**. However, it predominates later in states that have endured a severe third wave relative to states with current lower SARS-CoV-2 prevalence **(Fig 1d)**.

### No fourth wave of infection in the absence of new contagious variants

We performed equivalent analyses but assumed no introduction of variant B.1.1.7 into the population. Assuming a vaccine with high efficacy against infection (VE_SUSC_=90%, VE_SYMP_=10%, VE_INF_=10%) and national target vaccination rates (3500 per day) a fourth wave is projected to be mild or absent, and to not require further partial lockdown, even when the lowest threshold *C*_*max*_ is applied. In this scenario, cumulative incidence of infection by the end of 2021 is considerably lower and determined only by the extent of the third wave **(Fig 1c)**. Under King County conditions **(Fig 1c, orange line)**, we project cumulative infections and deaths to be 398,608 (18%) and 1512 (0.07%) the end of 2021.

### No difference in the magnitude of the fourth wave of COVID-19 infections and death due to changes in case threshold for relaxing partial lockdowns

*C*_*min*_ had limited influence on infections, hospitalizations and deaths in 2021 **(Fig 1a**). Lowering *C*_*min*_ did delay onset of the 4^th^ wave and the need for partial lockdown by approximately a month (not shown). This delay may prove valuable if vaccination rates increase dramatically during the spring and summer.

### Reduction in COVID-19 cases and deaths, and days under partial lockdown due to high vaccination rate

Across all scenarios, vaccination rate had a strong negative correlation with infections, hospitalizations and death, as well as days at maximal social distancing **(Fig 2a**). Given a vaccine with strong preventative efficacy (VE_SUSC_=90%, VE_SYMP_=10%, VE_INF_=10%) and currently anticipated triggers for partial lockdown (*C*_*max*_ *=* 350), increasing vaccination rate to 5000, 8000 or 11000 per day (0.23%, 0.36% and 0.46% of the population per day) would substantially decelerate the expansion of a fourth wave in the spring of 2021 **(Fig 2b)** resulting in 616,676 (28%), 633,145 (29%) and 581,166 (27%) projected total infections, and 1753 (0.08%), 1766 (0.08%) and 1667 (0.076%) projected total deaths respectively. 5000 vaccination per day would decrease the period of required lockdown during summer 2021 relative to 2000 vaccinations per day. 8000 vaccinations per day would blunt and delay the peak of the fourth wave until late 2021: this would eliminate the need for further partial lockdown, though lack of lockdown would allow slightly more infections by 2022 as a result.

**Figure 2.**
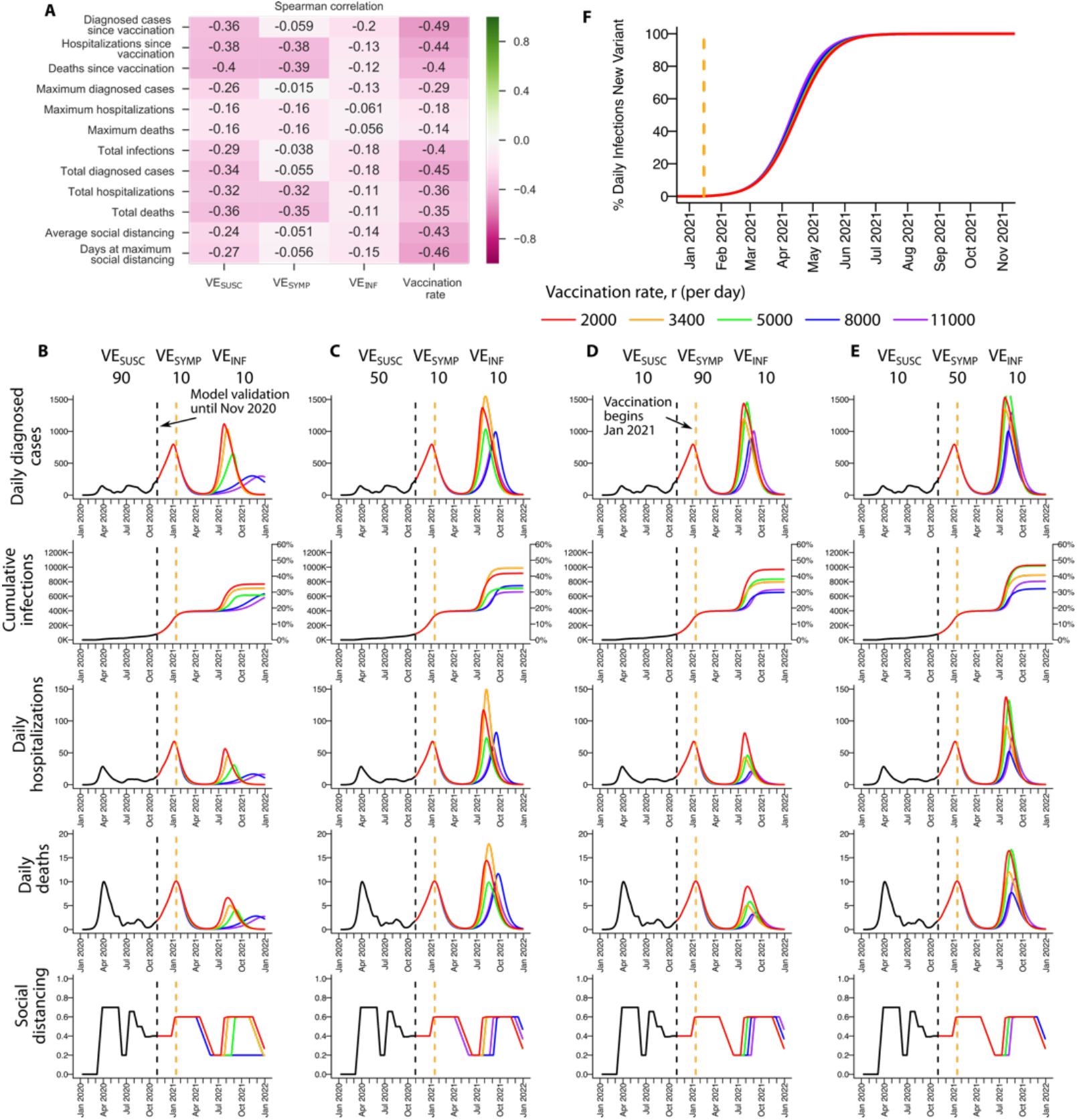
Increasing vaccination rate minimizes infections, hospitalizations and deaths, as well as total numbers of days under lockdown. **a**. Spearman correlations for input variables related to vaccination rate and efficacy. Increasing vaccination rate reduces infections, hospitalizations and deaths, and time under partial lockdown. Increasing vaccine efficacy against infection (VE_SUSC_) results in uniform benefit for all health-related outcomes and slight reduction in days at maximal social distancing. Increasing vaccine efficacy against symptoms given infection (VE_SYMP_) results in decreases in hospitalizations and deaths. Increasing vaccine efficacy against infectivity given infection (VE_INF_) results in slight reduction in numbers of diagnosed cases and infections. **b-e**. Impact of vaccination rate on relevant outcomes assuming different efficacy. **b**. VE_SUSC_=90% / VE_SYMP_=10% / VE_INF_=10%, **c**. VE_SUSC_=50% / VE_SYMP_=10% / VE_INF_=10%, **d**. VE_SUSC_=10% / VE_SYMP_=90% / VE_INF_=10%, **e**. VE_SUSC_=10% / VE_SYMP_=50% / VE_INF_=10%. All simulations *C*_*max*_ =350 cases per 100,000 (2-week average), *C*_*min*_ =25 cases per 100,000 (2-week average) and *σ*_*min*_ *=*0.2. Black dashed vertical line is the end of model calibration. Orange dashed vertical line is the time at which vaccination is initiated. **f**. Proportion of prevalent infections in **b**. due to new variants.

A vaccine with less efficacy but the same mechanism (VE_SUSC_=50%, VE_SYMP_=10%, VE_INF_=10%) would have comparable relative improvements at higher vaccination rates but would save fewer lives and prevent fewer infections **(Fig 2c)**. Under scenarios most compatible with King county, cumulative infections would increase from 689,057 (31%) **(Fig 2b, orange line)** to 969,161 (44%) **(Fig 2c, orange line)** while cumulative deaths would increase from 1806 (0.08%) to 2565 (0.12%). Recurrent partial lockdown is projected at all vaccination rates.

Similar vaccination-rate-dependent reductions in hospitalization and deaths were projected assuming vaccines that prevented symptoms rather than infection among infected people (VE_SUSC_=10%, VE_SYMP_=90%, VE_INF_=10%: **Fig 2d**, VE_SUSC_=10%, VE_SYMP_=50%, VE_INF_=10%: **Fig 2e**), though numbers of cumulative infections were considerably higher due to limited protection against secondary transmission. Under VE_SYMP_=90% scenarios most compatible with King county, cumulative infections and deaths are projected to be 785,870 (36%) and 1840 (0.084%) **(Fig 2d, orange line)**. Recurrent partial lockdown is projected at all vaccination rates.

For each scenario, strain replacement with the B.1.1.7 variant occurred rapidly during the fourth wave between March and June of 2021 **(Fig 2f)**.

### Reduction in COVID-19 cases and deaths, as well as time under partial lockdown due to higher vaccine efficacy against infection

We previously demonstrated that vaccines which prevent transmission, either by preventing infection (VE_SUSC_), or by preventing secondary transmission despite infection (VE_INF_), will decrease infections and deaths.^31^ With vaccines that only reduce symptoms in infected people (VE_SYMP_), achieving rapid herd immunity is contingent on concurrent high VE_INF_. In our current simulations with the B.1.1.7 variant, VE_SUSC_ negatively correlates with infections, hospitalizations, deaths and number of days under partial lockdown; VE_SYMP_ negatively correlates with total hospitalization and deaths; VE_INF_ slightly negatively correlates with total infections and diagnosed cases **(Fig 2a)**. However high efficacy against infection is not sufficient to prevent a fourth wave or high numbers of deaths if *C*_*max*_ is high **(Fig 1b)** or vaccination rate is low **(Fig 2b)**.

### Low case threshold for triggering partial lockdown and high vaccination rate: essential variables to limit SARS-CoV-2 infections and deaths

We include contour plots in which more granular sampling across included ranges of vaccination rates and lockdown thresholds were performed at four fixed vaccine scenarios, two of which would be compatible with results from Moderna and Pfizer trials (VE_SUSC_=90%, VE_SYMP_=10%, VE_INF_=10%: **Fig 3a** or VE_SUSC_=10%, VE_SYMP_=90%, VE_INF_=10%: **Fig 3b**) and two of which anticipate the possibility of less efficacy after a single dose of these vaccines and/or less efficacy against novel variants (VE_SUSC_=50%, VE_SYMP_=10%, VE_INF_=10%: **Fig 3c** or VE_SUSC_=10%, VE_SYMP_=50%, VE_INF_=10%: **Fig 3d**). These scenarios capture vaccines which either effectively limit **(Fig 3a, c)** or do not effective limit **(Fig 3b, d)** secondary transmission from vaccine recipients.

**Figure 3.**
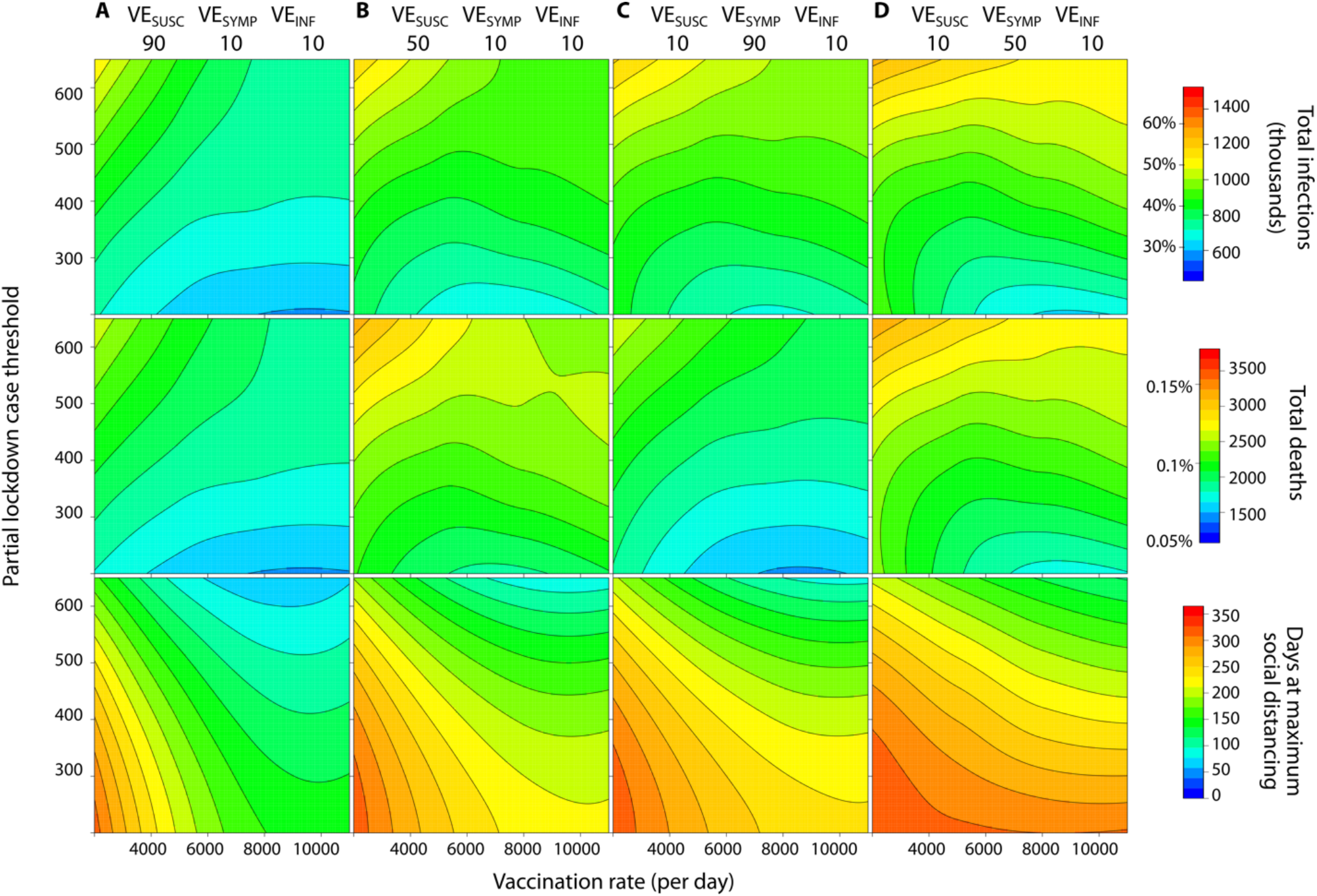
High vaccination rate and low case threshold for triggering partial lockdown are the key variables for limiting total infections and deaths regardless of vaccine efficacy profile. Heat maps demonstrating joint effects of vaccination rate (x-axis) and case threshold for triggering partial lockdown (y-axis) are shown for four plausible vaccine profiles. **a**. VE_SUSC_=90% / VE_SYMP_=10% / VE_INF_=10%, **b**. VE_SUSC_=50% / VE_SYMP_=10% / VE_INF_=10%, **c**. VE_SUSC_=10% / VE_SYMP_=90% / VE_INF_=10%, **d**. VE_SUSC_=10% / VE_SYMP_=50% / VE_INF_=10%. Outcomes are total infections (top row), total deaths (middle row) and days under partial lockdown after vaccination initiation (bottom row). Increasing vaccination rate lowers infections and deaths across all scenarios. Increasing vaccination rate substantially lowers total days under lockdown, particularly when case threshold trigger for partial lockdown is low. Lowering case thresholds for triggering partial lockdown decreases total infections and deaths but results in higher number of days under partial lockdown in many scenarios. A decrease in VE_SUSC_ (**a** to **b**) results in more infections and deaths with only slight impact on time under lockdown. A decrease in VE_SYMP_ (**c** to **d**) results in more infections and deaths with only slight impact on time under lockdown. VE_SUSC_ provides a substantial reduction in infections but slight reduction in deaths relative to an equivalent VE_SYMP_ (**a** to **c** and **c** to **d**). All scenarios assume *C*_*min*_ =25 cases per 100,000 (2-week average) for relaxing partial lockdown and *σ*_*max*_ *=*0.2.

Key findings are that 1) decreasing *C*_*max*_ by several hundred cases may lower total infections during 2021 by several hundred thousand and total deaths by several hundred, 2) decreasing *C*_*max*_ by several hundred cases may increase total days under lockdown during 2021 by over a month (though approximately 100 of the total reported days under partial lockdown have already been accrued during the third ongoing wave), 3) increasing the vaccination rate by several thousand people per day may lower total infections by more than one hundred thousand, total deaths by more than one hundred, and days under lockdown by two months, 4) increasing the vaccination rate lowers cases and deaths under all scenarios, 5) increasing the vaccination rate lowers total days under partial lockdown most significantly when *C*_*max*_ is low, 6) while equivalent trends are noted across all four vaccine efficacies, high VE_SUSC_ rather than VE_SYMP_ results in fewer infections but only slightly fewer deaths at equivalent *C*_*max*_ and vaccination rate, and 7) higher vaccine efficacy (either against infection or symptoms) also results in fewer infections and deaths at equivalent *C*_*max*_ and vaccination rates.

### Importance of high vaccination rate given viral variants with greater infectivity or more frequent introduction of these variants into King County

We performed equivalent analyses but varied degrees of variant infectivity. With only a 35% increase in infectivity, increasing vaccination rate above 8000 per day prevented a fourth wave of infections and deaths **(Fig 4a)**. Increasing variant infectivity to 75% resulted in a more severe fourth wave which could be dampened but not prevented with more rapid vaccination **(Fig 4b)**. However, increasing the rate above 8000 per day lowered the time required under partial lockdown in this scenario.

**Figure 4.**
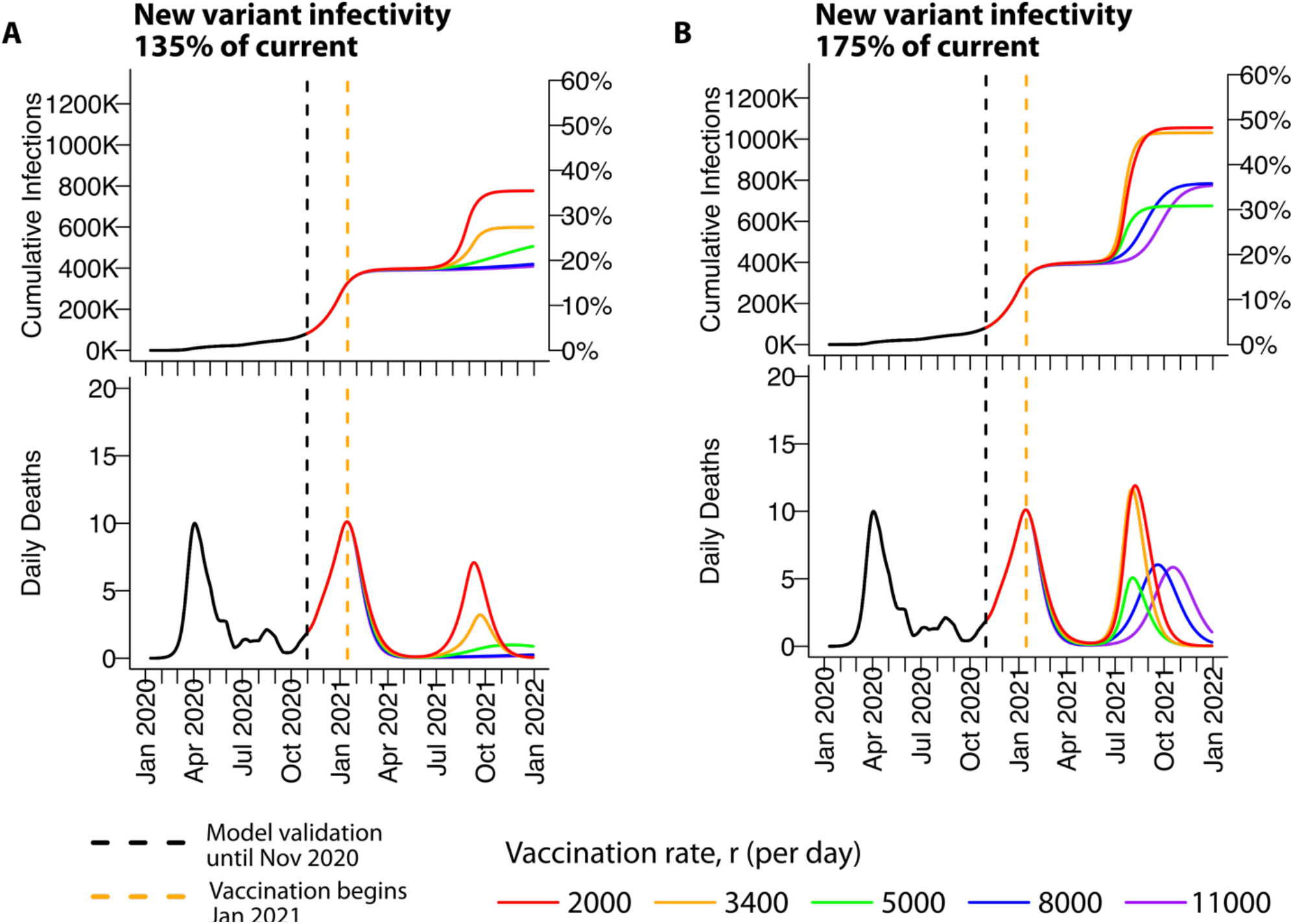
High vaccination rate, limits infections and deaths with either 35% or 75% more infectious novel SARS-CoV-2 variants. Impact of varying vaccination rate on relevant outcomes. **a**. A 35% more infectious variant is assumed. High enough vaccination rate can prevent a fourth wave of infections and deaths. **b**. A 75% more infectious variant is assumed. Higher vaccination rate delays the fourth wave and lowers cumulative infection and peak death. All simulations assume VE_SUSC_=90% / VE_SYMP_=10% / VE_INF_=10%,*C*_*max*_ =350. and *C*_*min*_ =25 and *σ*_*min*_ *=*0.2.

We next increased the daily introduction rate of contagious variants into King County **(Sup fig 4)**, which only slightly increased the intensity of the fourth wave of cases and did not impact the critical importance of high vaccination rate.

### Importance of high vaccine uptake to achieve benefits of high vaccination rate

We performed equivalent analyses but decreased or increased the percent of people in each age cohort receiving a vaccine. Achieving 100% vaccine uptake would substantially limit the severity of fourth waves particularly with vaccination rates exceeding 8000 per day **(Sup fig 5a)**. However, if uptake decreased to 60%, higher peaks in daily deaths are projected during the fourth wave and the benefit of rapid vaccination is almost completely lost **(Sup fig 5b)**.

### Importance of maintaining moderate social distancing during vaccine roll out and between third and fourth waves of infection

We performed equivalent analyses but increased **(Sup fig 6a)** or decreased **(Sup fig 6b)** the degree of social distancing when partial lockdown was relaxed. We again noted that increased vaccination decreased cases and deaths under both scenarios. Increasing the minimum social distancing maintained between waves of infection substantially decreased total infections and allowed prevention of a fourth wave assuming high vaccination rate **(Sup fig 6a)**.

## Discussion

It is often stated that vaccines do not save lives, vaccinations do. Our modeling strongly reinforces this point. High efficacy of vaccines against new more contagious variants, and in particular their ability to block ongoing transmission rather than simply preventing symptoms, will potentially prevent thousands of infections and save hundreds of lives in King County.^31^ However, the vaccination rate is of even greater importance and will determine the extent of a fourth wave in the spring and summer of 2021 for all plausible vaccine efficacies. This wave has the potential to be particularly lethal, given that a more contagious variant will predominate, but may be quite limited if 8000 people (0.36% or 1 in 280 people) or more can be vaccinated daily.

Rapid vaccination will have the greatest absolute benefit in states such as Hawaii, Vermont, Washington and Oregon that have maintained lower overall seroprevalence to date. These states often employed more rapid triggering of partial lockdown during recent waves of SARS-CoV-2 infection. Because a higher proportion of people are still susceptible in these states, they are particularly vulnerable to contagious variant strains. A larger percentage of people will in turn need to be vaccinated to reach herd immunity. Aside from lowering infections, hospitalizations and deaths, more rapid vaccination will also decrease the total number of days under partial lockdown.

Our model projects that with the new more infectious variants, higher case thresholds for triggering partial lockdown will result in greater numbers of total infections and deaths per capita.^18^ Even with the current consensus variant, this projection has been observed during prior waves of infection: multiple states in the northern plains and southwest recently endured a severe third wave relative to Washington state. In these states, widespread vaccination may contribute relatively less to generation of herd immunity, which instead will be reached in part from a high number of cases. This conclusion is most relevant if existing vaccines retain efficacy for both variants and if prior infection is cross-protective against new variants. Some concern for waning immunity or lack of cross-variant protection exists based on recent experience in Manaus where a severe wave of infection is ongoing despite high pre-existing seroprevalence.^42^

Our simulations suggest that lowering the case threshold for partial lockdown will most significantly decrease the number of total infections in 2021 if coupled with a high vaccination rate. This is because the high infectivity of B.1.1.7 and other emerging variants will make fourth waves severe, even if partial lockdown is initiated relatively early. The current harrowing experiences in London and Manaus are consistent this projection. Lowering the case threshold for partial lockdown will decrease total deaths under all scenarios assuming high efficacy vaccines because this strategy will allow more time to vaccinate the elderly.

We demonstrate that increasing physical distancing and masking between third and fourth waves (increasing minimum SD in the model) will also decrease cases and deaths. Strategies for pandemic management during 2021, both during and between waves, should mirror and improve upon those taken during the early months of the pandemic: widespread masking with N95 masks in crowded environments,^19^ avoidance of large crowds, and a focus on improved ventilation in high-risk indoor environments. Prevalent use of rapid, inexpensive testing could aid in efficient contact tracing and case “cluster busting”.^20^ Yet, these measures are likely to be insufficient if known super-spreader environments such as crowded restaurants and bars are also not closed for a short period of time. Unfortunately, lowering the case threshold for partial lockdown may result in a slightly longer time under lockdown overall in 2021, though some of this time has already been accrued.

Importantly, the key drivers of health-related outcomes hold under various vaccine profiles including lower overall vaccine efficacy, or a shift from protection against infection to protection against symptoms, both of which are possible with emerging new variants. These scenarios would lead to more overall infections and deaths, and would only add urgency to the need to vaccinate as quickly as possible and set low case thresholds for partial lockdown. We also demonstrate that vaccine uptake is crucial. If only 60% of the elderly cohort accepts a vaccine, then a severe fourth wave with requirement for partial lockdown is likely.

A similar conclusion can be made for variants that increase infectivity by 75%, as appears to be possible with the South African B.1351 lineage.^16^ In this scenario, avoidance of a fourth wave may unfortunately not be possible. However, rapid vaccination will blunt and delay the fourth wave, perhaps avoiding the requirement for partial lockdown. Maintaining high levels of masking and physical distancing between waves is crucial under this circumstance.

The current Pfizer and Moderna mRNA vaccines have extremely high efficacy against disease but require two doses.^35,36^ Initial data from the Moderna trial suggests the possibility of >50% protection against asymptomatic infection following the first dose, though confirmation is required in studies not including a second dose. The Johnson and Johnson vaccine is 69% effective and is given as a single dose.^38^ Our results highlight the critical importance of evaluating single dose regimens because doubling the rate of vaccination assuming equivalent efficacy could prevent thousands of infections, hundreds of deaths and dozens of days under partial lockdown in King County alone and could also be the factor which eliminates a meaningful surge of cases in spring 2021.

By simulating the model without introducing B.1.1.7, we provide a negative control to assess the additional damage that is expected due its higher infectivity. Unfortunately, the new variant is likely to contribute to excess infections and deaths. Vaccination scenarios which would have achieved herd immunity quickly enough to prevent a fourth wave with the current dominant variant will fail to do so with B.1.1.7. The timing and severity of the fourth wave is nevertheless difficult to project based on uncertainty of variant infectivity, current prevalence of contagious variants in the community, possible loss of vaccine efficacy against new variants and uncertainty regarding adherence to masking and physical distancing in the upcoming months.

While our model is calibrated closely to King County Washington, it allows various case thresholds for partial lockdown and distribution rates. To this end, its projections are relevant for regions with lower and higher seroprevalence, and it is useful for understanding future different case trajectories in multiple states.

However, several limitations are notable. Our contact network is stratified by age and not race or profession and therefore is likely to miss important heterogeneities in transmission risk. We only assume initial vaccine allocation predominately to the elderly and a subset of younger health care workers to match current plans. While we make general projections about single versus double vaccination, we do not consider the specifics of vaccine scheduling and delays in accrued protection against infection.

We also assume no waning immunity following natural infection or vaccination, and do not include the possibility of added protection in people who are first infected and then vaccinated. Based on existing data demonstrating prolonged antibody response after vaccination and natural infection,^43,44^ we believe that this issue can be neglected over the time intervals we model. Rapid loss of immunity would enhance the need to vaccinate quickly. Careful surveillance for SARS-CoV-2 re-infection, as well as viral antigenic drift, is needed urgently and will dictate the need for re-vaccination. Rapid institution of partial lockdown and vaccination will remain the two key strategies to limit deaths and number of days under lockdown.

Finally, we project a 2 to 3-month delay until new variant cases surge. This is based on the fact that among sequenced viruses in the United States, new variants appear relatively uncommon at this stage representing fewer than 0.5% of sequenced viruses.^18^ However, if current estimates are misleadingly low, then a wave of infections can be expected sooner when a lower proportion of the population has been vaccinated and/or infected.

In conclusion, we project that despite considerable uncertainty about the timing and severity of variant waves in the United States, rapid vaccine distribution and low case threshold for triggering partial lockdown are the two critical components to save the most lives.

## Methods

### King Country transmission model

We modified a previously developed deterministic compartment model,^45^ which captures the epidemic dynamics in King County, WA between January 2020 and October 2020 and projects the trajectory of the local pandemic through the end of 2021 in the absence and presence of vaccines^31^. Vaccination is simulated with a starting date of January 1, 2021. We include the possibility of infection with a new variant with a potentially higher infectivity, which is assumed to already be circulating at undetectable levels and is also imported continuously throughout the coming months.

Our mathematical model is an extension of an SIR model in which each state variable (or compartment) can be stratified by age (4 compartments, children <20, young adults 20-40, adults 40-70, and seniors >70), vaccine status (yes or no), and infecting strain (current or B117 strain). Thus, state variables are tensors 4(*a, V, q*)with dimensions for age group, vaccine status, and infecting strain. We include compartments for susceptible, exposed, pre-symptomatic, asymptomatic, symptomatic, hospitalized, and deceased. We also include separate compartments for diagnosed cases of each infected type both for the original model fitting to cases but also because diagnosed cases may behave differently from individuals who do not know their infection status. The model schematic and illustrations of case-dependent social distancing and vaccine roll-out are shown in **Supp fig 1**. All mathematical details are presented in the **Supplementary Information**.

### Modeling vaccination and case-dependent partial lockdowns

Vaccination is modeled as a constant distribution of vaccines + (doses per day) that are given first to at 80% to seniors (70+) and 20% to adults. Children are not vaccinated. When a maximum coverage (*V*_*max*_) is reached in the senior group, distribution continues to adults.

Vaccines can work by several mechanisms simultaneously. They can completely block infection (VE_SUSC_ – reducing the number of susceptible individuals), block symptomatic disease (VE_SYMP_ – adjust the fraction of infections that are symptomatic vs asymptomatic), and/or decrease the possibility of onward transmission after infection (VE_INF_ – reducing the force of infection). Each vaccine efficacy ranges from 0-1.

To attempt to accurately model societal or governmental responses to pandemic surges, we include a time-dependent term that reduces contacts when cases reach a certain point. This term *σ* (*a, t*)varies from 0, indicating pre-pandemic levels of societal interactivity and no masking, to 1, indicating complete lockdown with no interactions **(Supp Fig 1b)**. When two-week number of diagnosed cases per 100,000 people rises above *C*_*max*_, a “partial-lockdown” (*σ* (*a, t*)→ *σ*_*pL*_)is mandated **(Supp Fig 1b)**. Partial lockdown is defined by an enforced social distancing of *σ*_*pL*_ =0.6 in the 3 younger age cohorts and *σ*_*pL*_ =0.8 in seniors (≥70 years). When cases drop below a relaxation threshold *C*_*min*_ *σ* (*a, t*)is lowered gradually from *σ*_*pL*_ to *σ*_*min*_(*a*) at a rate of 10% reduction in *σ* (*a, t*) every two weeks that cases stay below the threshold. Notably, minimum distancing of seniors is still 20% higher than in younger cohorts.

Several other explored parameters (see **Supp figs 4-7** include the infectivity of the new variant, the rate of imports of the new strain, and variation in minimal social distancing.

### Model simulation

Ordinary differential equations are used to simulate the model using *lsoda* in R.

### Model calibration and parameterization

Model calibration and parameterization was performed in Ref. ^31^. This involved fitting the model to Diagnosed Cases, Hospitalizations, and Deaths from Department of Health data in King County Washington. Age proportions and age structured contact matrices were inferred from local demographic and contact data. All initial conditions and parameters for the model can be found in **Sup Table 2**.

### Global sensitivity analysis

We performed a comprehensive exploration of vaccination rate, mechanisms, and case levels for triggering and releasing lockdown: all combinations of all parameters in Table 1. 12 outcomes quantifying societal and public health were recorded (see **Sup fig 2**) for each simulation. Then, correlation and clustering analyses were performed by calculating Spearman correlations among outcomes and between variables and outcomes using the Seaborn package in Python.

### Code

All R code for model simulations are freely available at www.github.com/FredHutch/COVID_modeling. Custom plotting scripts in R and Python are available upon request.

## Data Availability

All code is available as described in the supplementary information.

---

**Supplementary figure 1.**
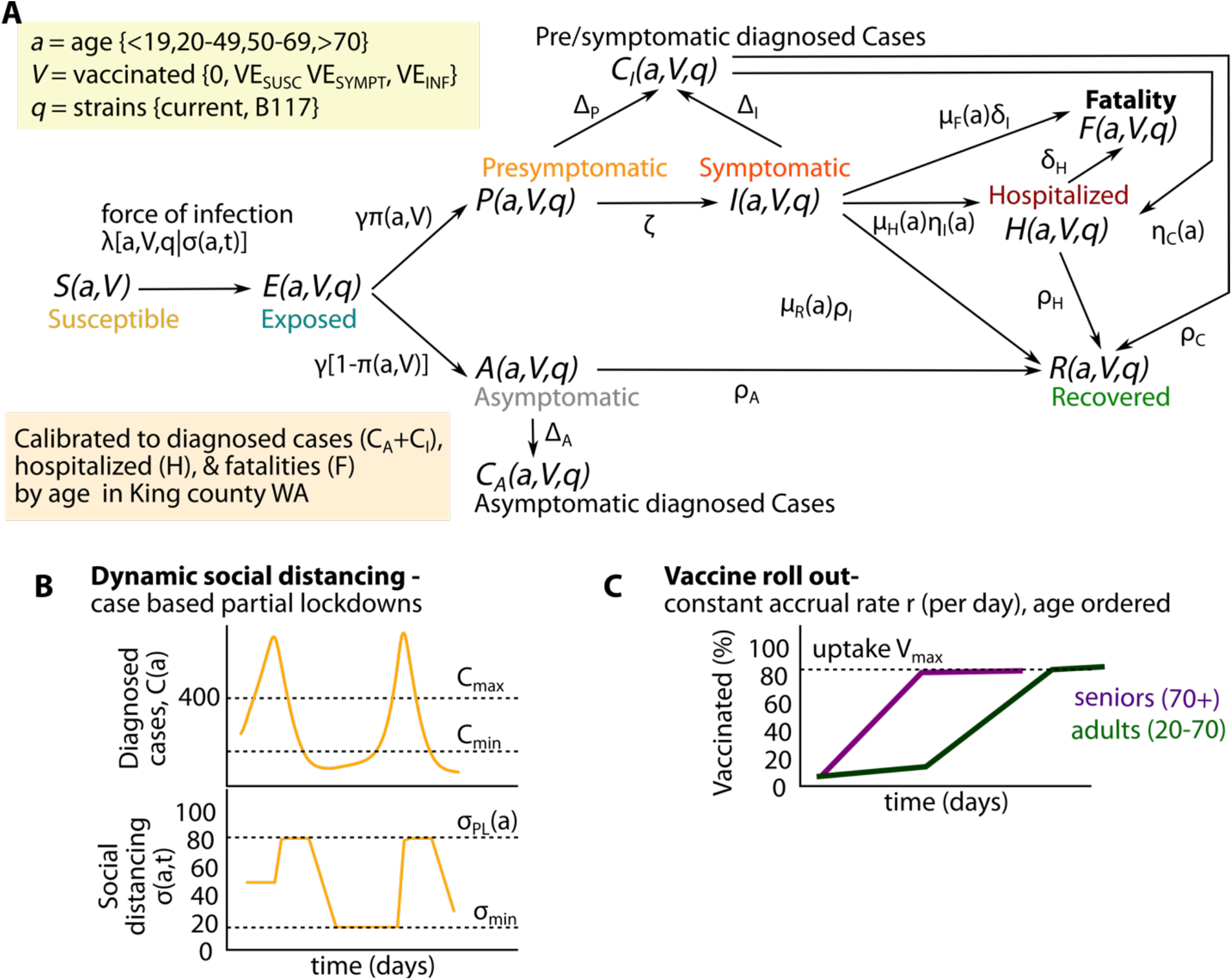
Mathematical model schematic. **a**. Model structure, **b**. Dynamic social distancing framework, **c**. Vaccination rollout plan.

**Supplementary figure 2.**
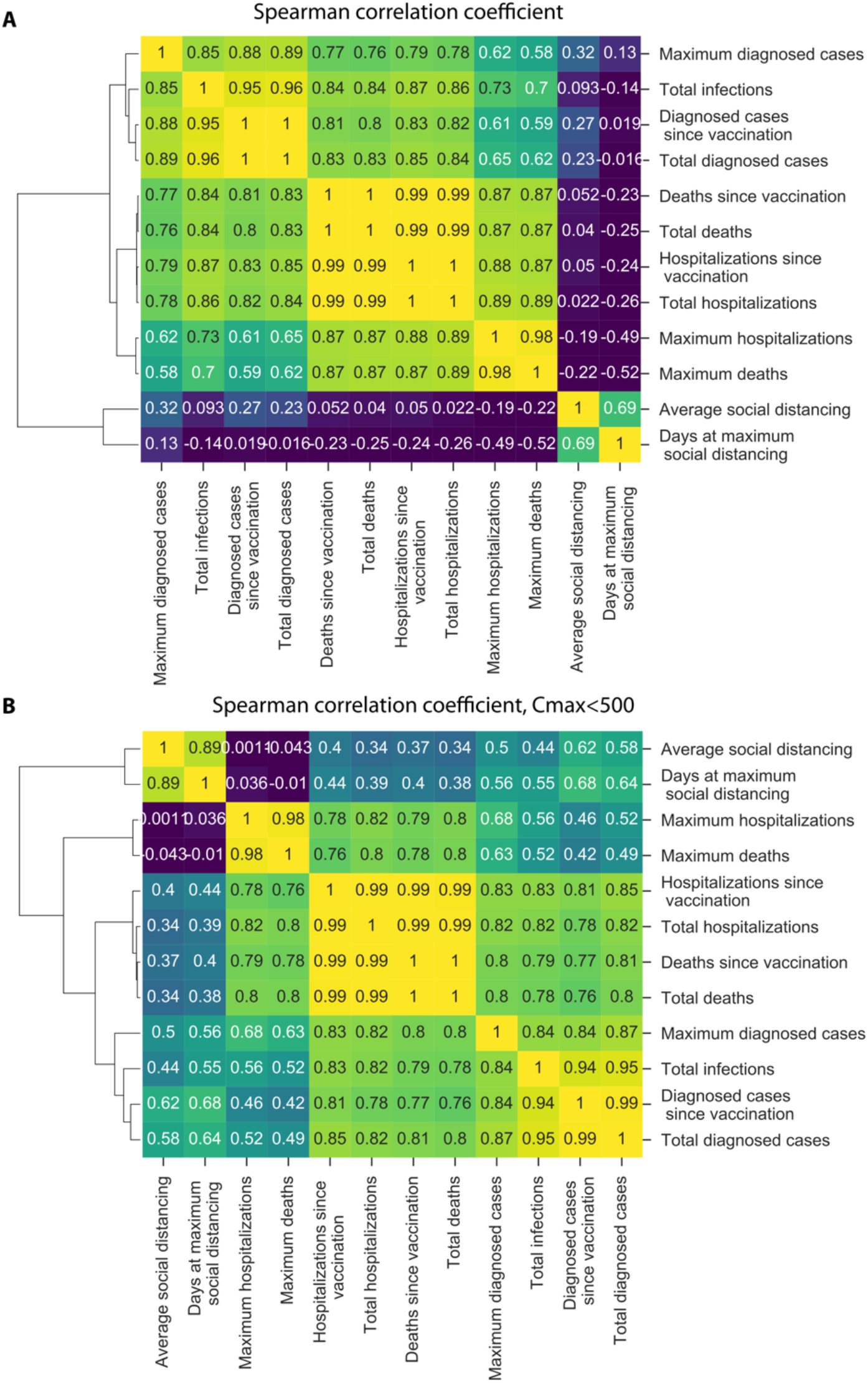
Correlations among model outcomes. **a**. Spearman correlation coefficients between model outcomes from 3888 simulations, **b**. Spearman correlation coefficients between model outcomes excluding all *C*_*max*_ =650 scenarios where partial lockdown does not occur during the third wave of infection.

**Supplementary figure 3.**
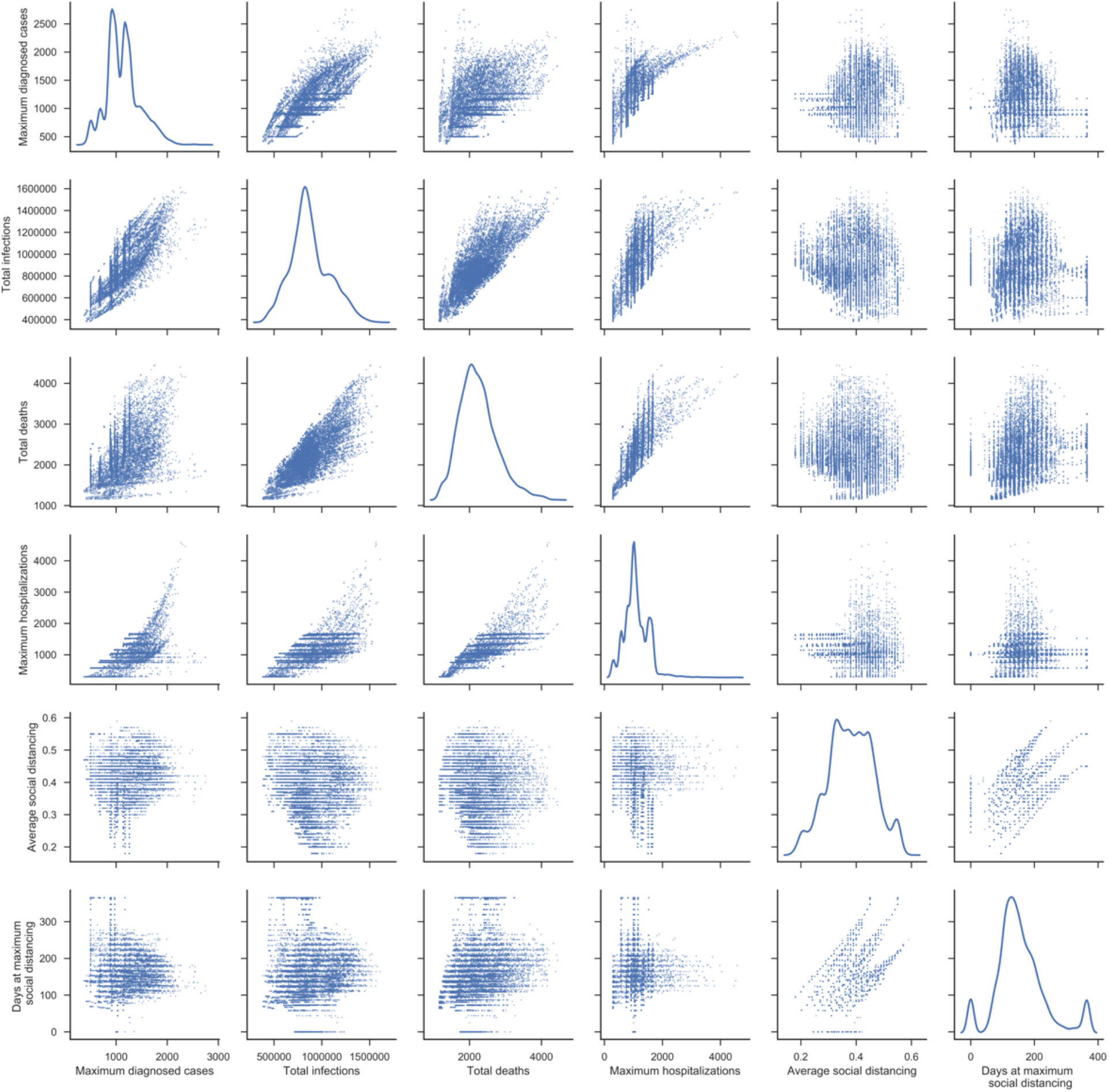
Individual simulations with projections of outcomes. Dot plots represent relationship between variables listed on the x- and y-axes. Each dot represents a single simulation with a unique parameter set. Line diagrams represent density plots for a given outcome projected on the x-axis.

**Sup fig 4.**
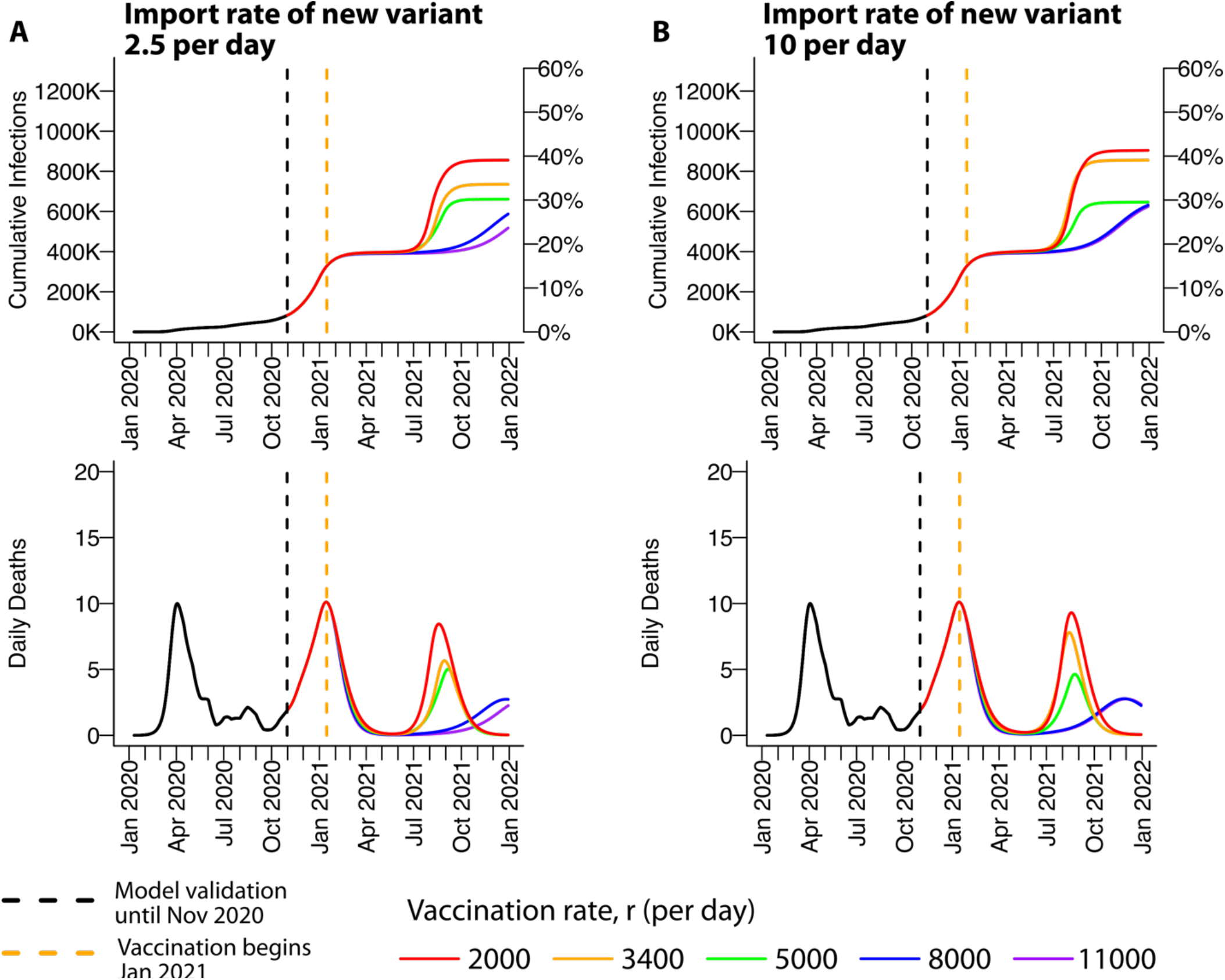
High vaccination rate limits infections and deaths with slow or rapid introduction of a 55% more infectious SARS-CoV-2 variant into King County. **a**. An import rate of 2.5 cases per day is assumed. **b**. An import rate of 10 cases per day is assumed. In both scenarios, a higher vaccination rate delays the fourth wave and lowers cumulative infection and peak death. All simulations assume VE_SUSC_=90% / VE_SYMP_=10% / VE_INF_=10%, *C*_*max*_ =350. and *C*_*min*_ =25.

**Sup fig 5.**
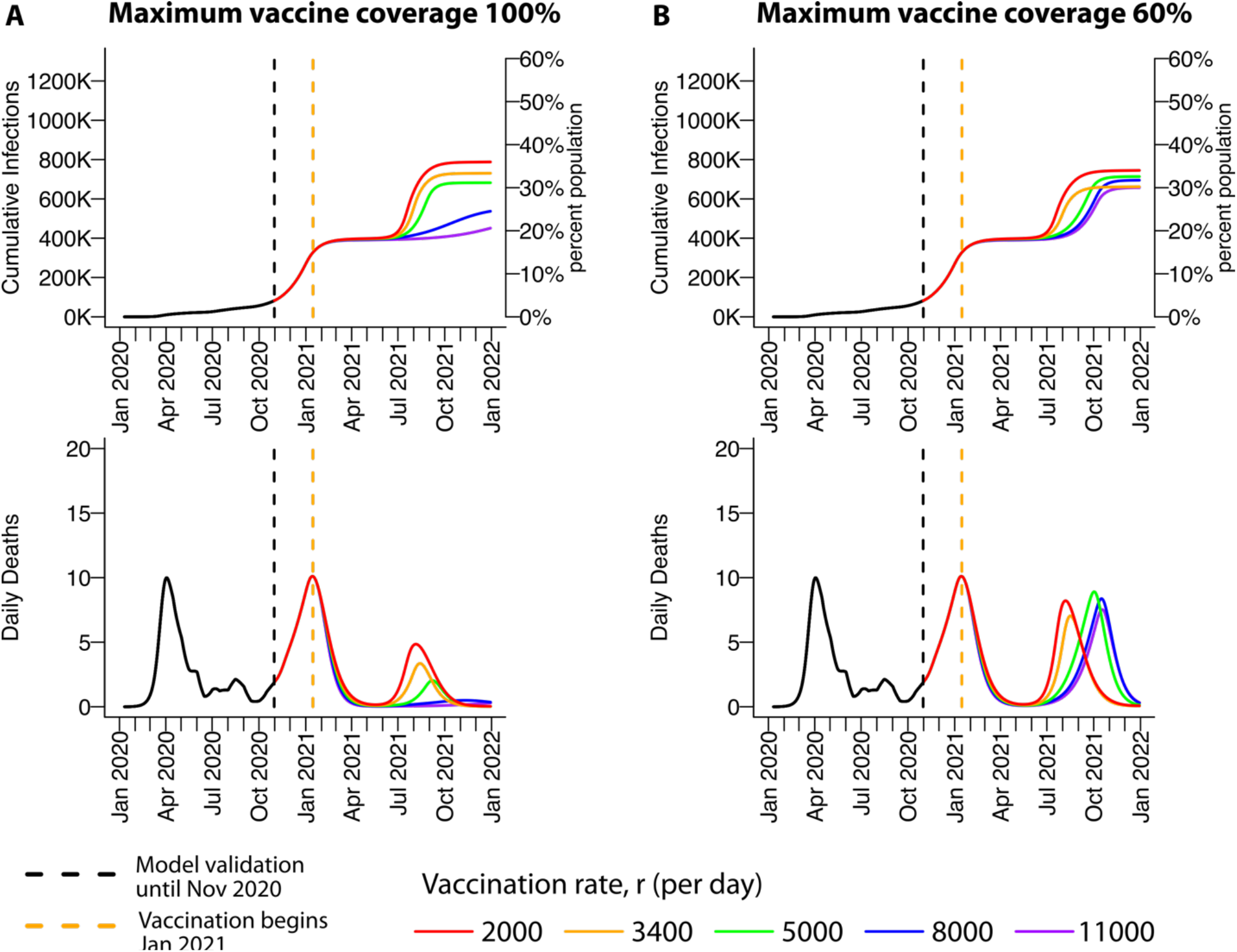
High vaccination rate limits infections and deaths only with sufficient vaccine uptake. Impact of varying vaccination rate on relevant outcomes. **a**. 100% of each age cohort is assumed to receive the vaccine. High enough vaccination rate can prevent a fourth wave. Higher vaccination rate prevents a large fourth wave **b**. 60% of each age cohort is assumed to receive the vaccine. Higher vaccination rate delays but does not blunt the fourth wave and lowers cumulative infection and peak death. All simulations assume VE_SUSC_=90% / VE_SYMP_=10% / VE_INF_=10%, *C*_*max*_=350 and *C*_*min*_ =25.

**Sup fig 6.**
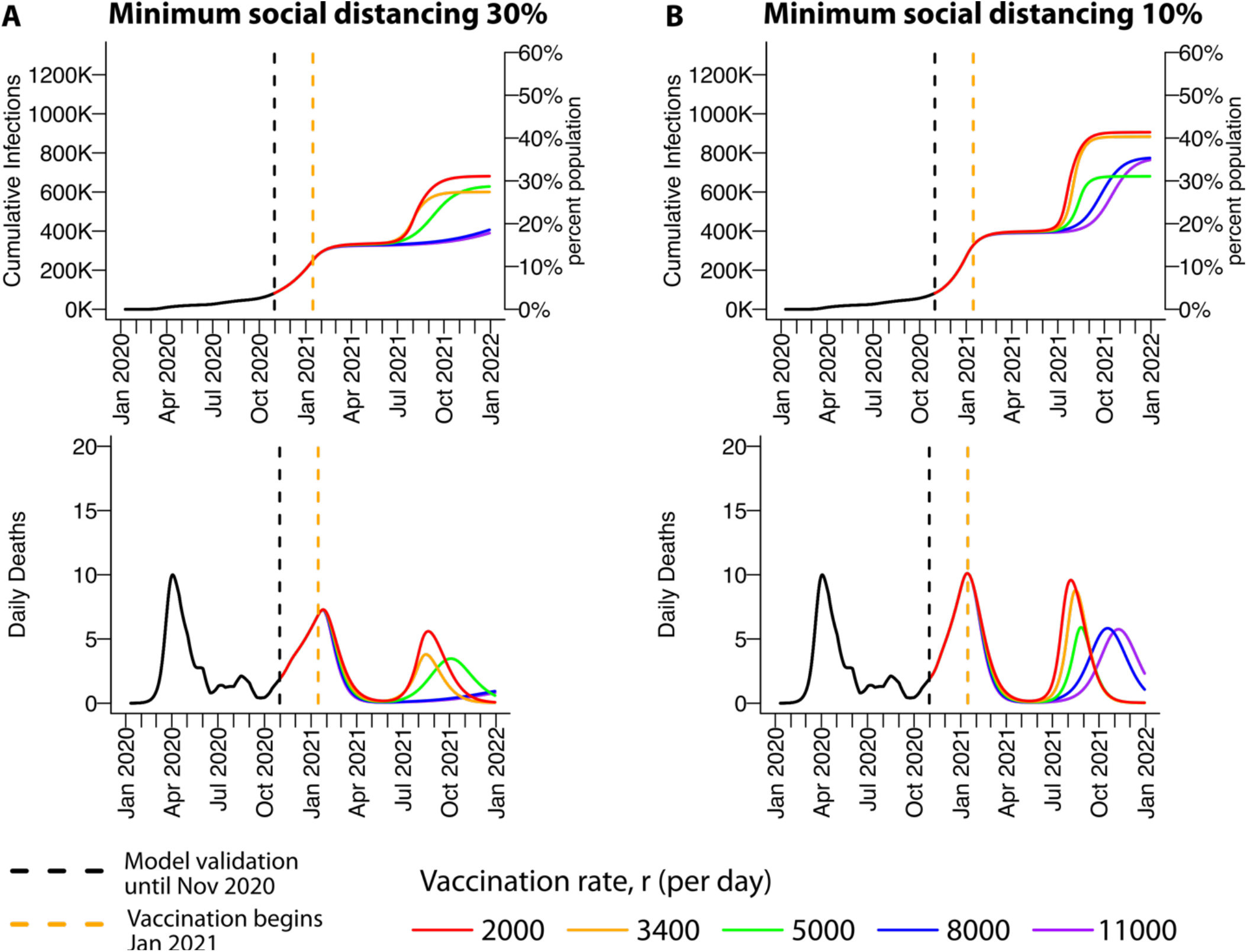
High vaccination rate limits infections and deaths to a greater degree when higher social distancing is maintained between partial lockdown periods. Impact of varying vaccination rate on relevant outcomes. **a**. A minimum 30% social distancing value is assumed. High enough vaccination rate can almost prevent a fourth wave. **b**. A minimum 10% social distancing value is assumed. Higher vaccination rate delays but does not prevent the fourth wave and lowers cumulative infection and peak death. All simulations assume VE_SUSC_=90% / VE_SYMP_=10% / VE_INF_=10%, *C*_*max*_ =350. and *C*_*min*_ =25.

## Supplementary Information: COVID SA

In this supplementary information we include details of the extended SIR model. All code is freely available at www.github.com/FredHutch/COVID_modeling.

To model SARS-CoV-2 epidemiological dynamics we have applied a dynamical systems approach which uses an SIR model extended in several key ways. In general each state variable is now a tensor whose dynamics are given by its properties, distinguished as age group *a*, vaccination status *V*, and depending on the infecting strain *q*. The model extensions are detailed in Table S1 and a schematic cartoon is provided in Fig S1.

**Table S1:**
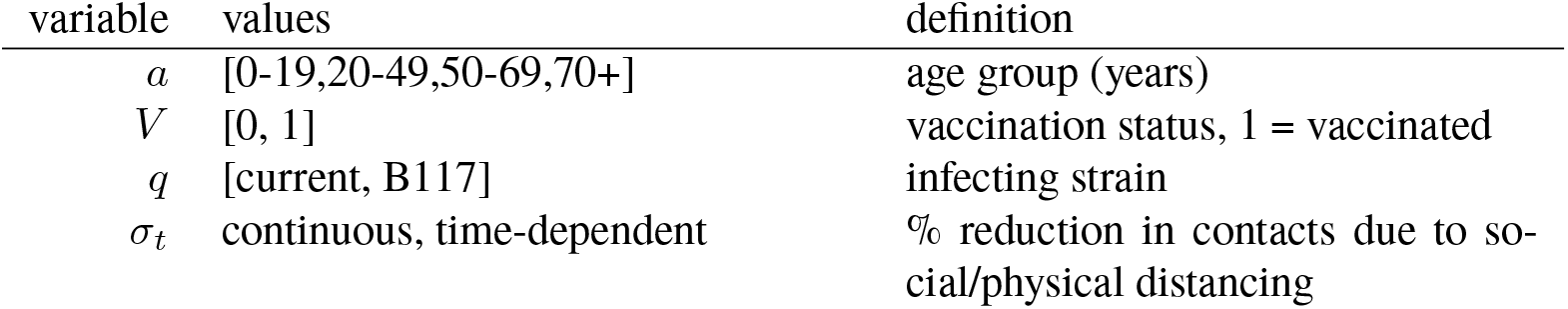
Variables describing extensions to SIR model.

### Force of infection

Perhaps the most important equation for the model summarizing the dynamics is the ‘force of infection’. Here we show the value of the force of infection for each subset of state variables. The force of infection depends on the state of the infected individual (*X*) and the strain-specific infectivity *β*_*X*_(*q*). It also depends on a time-dependent reduction in contacts mediated by social/physical distancing *σ*_*t*_. Finally, we use the empirically derived contact matrix to adjust the force of infection on a certain age group from transmitters in each other age groups (denoted by *a*_*T*_, and calculated using the adjacency matrix *𝒜*(*a, a*_*T*_)). Finally, we have the force of infection for an individual in age group *a*, exposed to strain *q*, with vaccination status *V*, and with ongoing social distancing level *σ*_*t*_:

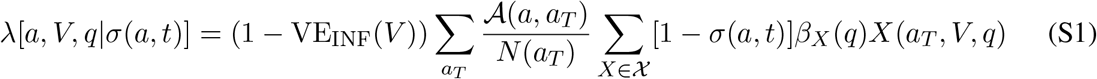

Where *𝒳* = *A, P, I, C*_*A*_, *C*_*I*_ is the set of all potentially infectious states. Note it is assumed that hospitalized individuals do not contribute to transmission (*β*_*H*_ = 0). Naturally, susceptible, exposed, recovered, and deceased individuals also do not contribute to ongoing infection.

The total number of individuals (across age, vaccination, and strain) in each compartment can then be calculated as a sum over the variables as

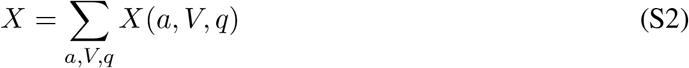

### Assumptions on asymptomatic infection

We assume that 20% of infections are asymptomatic and that asymptomatic people are as infectious as symptomatic individuals but missing the highly infectious pre-symptomatic phase. As a result, the relative infectiousness of individuals who never develop symptoms is 56% of the overall infectiousness of individuals who develop symptomatic COVID-19. This conservative estimate falls between the 35% relative infectiousness estimated in recent review based on 79 studies^1^ and the current best estimate of 75% suggested by the CDC in their COVID-19 pandemic planning scenarios.

### Dynamic social distancing

An attribute that sets our model apart from most others is a notion of dynamical social distancing related to the current diagnosed cases. We include a time-varying, age-stratified vector *σ* (*a, t*) that governs social distancing (non-pharmaceutical interventions) including reduced contacts through personal choices and/or mandated partial lockdowns, as well as reductions in exposure contacts due to mask wearing or physical distancing. *σ* (*a, t*) varies from 0, indicating pre-pandemic levels of societal interactivity and no masking, to 1, indicating complete lockdown with no interactions. This function is parameterized by 4 values: the maximum *C*_*max*_ and minimum *C*_*min*_ number of cases and the partial-lockdown and reopened social distancing values *σ*_*PL*_(*a*) and *σ*_*min*_(*a*) (see Supp Fig 1B). Thus we have

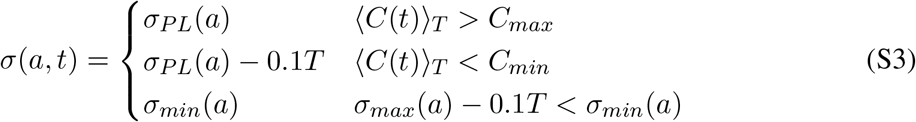

where the time average of cases ⟨*C*(*t*)⟩_*T*_ is taken over *T* = 2 week intervals in the current simulation. Thus, the system triggers lockdown if the average cases rises over the max threshold and distancing immediately becomes *σ*_*PL*_(*a*) which is 40% of prepandemic levels in non-seniors and 20% in seniors. Then, once cases drop below the release threshold *C*_*min*_, 10% of the distancing is removed every *T* weeks until reaching the minimum social distancing *σ*_*min*_(*a*). This value is not necessarily zero because we expect persistent features such as masking, work from home and avoidance of large social gatherings will continue to limit the number of interpersonal contacts relative to pre-pandemic levels.

### Vaccination mechanisms

The possibility of vaccination is further complicated by inclusion of 3 mechanisms. The vaccine can completely block infection (VE_SUSC_), adjust the fraction of infections that are symptomatic (VE_SYMP_), and/or decrease the possibility of onward transmission after infection (VE_INF_). Each vaccine efficacy ranges from 0-1.

The number of individuals in each age group *N* (*a*) is calculated at each step as

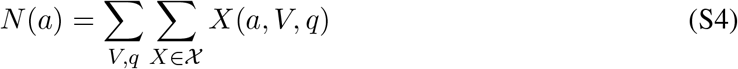

where 𝒳 = {*S, E, A, P, I, C*_*A*_, *C*_*I*_, *H, R*} is all non-deceased states. We also sum over the vaccinated and unvaccinated individuals, and over infecting strains.

### Vaccination program

The vaccination program is implemented to best-mimic the current practice of vaccinating mostly elderly first, and then adult age groups, but never children. Vaccination distribution follows a daily rate *r*. Thus we allow 80% of vaccines to go to elderly each day *S*(*a* = 70+,*V* = 1) = 0.8*rt* and the remaining 20% to adults *S*(20 *< a <* 70,*V* = 1) = 0.2*rt*. We set a maximum coverage *V*_*max*_ that roughly models vaccine uptake and compliance. Once the coverage is reached in the elderly, all vaccines are distributed to adults.

The whole set of equations thus is

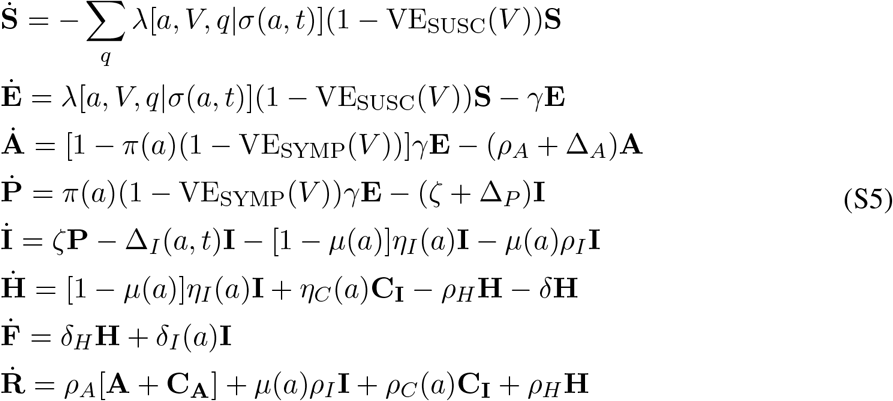

and equations for diagnosed cases follow

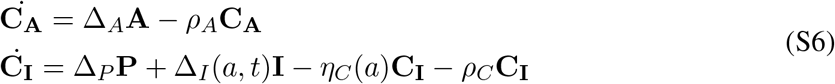

### Comments on diagnosed cases

In addition to the main set of differential equations (Eq. S5), we also track the equations that govern the accumulating diagnosed cases (*C*_*X*_, Eq. S6). Diagnosed cases can reenter the main equation set above because of hospitalizations due to diagnosed cases. We use the total diagnosed cases *C* = *C*_*A*_ + *C*_*I*_ to fit caseload data. A nuance is that diagnosis rates Δ(*a, t*) are explicitly time dependent, which is used to incorporate data on number of individuals in each age group that are seeking testing. Importantly, diagnosed individuals may behave differently, so they are separated in the model for this reason too.

### Initial conditions and model parameters

We use the state of the pandemic in October 2020 as derived in our prior publication ^2^ to initialize and parameterize the simulations in this manuscript.

Initial conditions are tabulated in Table S2. Fixed parameters are tabulated in Table S3, and estimated parameters are tabulated in Table S4.

---

**Table S2:** Initial conditions, no vaccinated individuals and no new strain, i.e. *V* =0 and *q*=current for all entries.

| State | Initial condition |  |  |  |
| --- | --- | --- | --- | --- |
|  | Children | Young Adults | Adults | Seniors |
| $S$ | 471690 | 950078 | 493341 | 169429 |
| $E$ | 1175 | 1973 | 925 | 297 |
| $A$ | 469 | 634 | 300 | 107 |
| $P$ | 624 | 932 | 423 | 139 |
| $I$ | 1763 | 2260 | 1139 | 509 |
| $C_A$ | 217 | 805 | 248 | 5 |
| $C_I$ | 43 | 158 | 46 | 0.73 |
| $H$ | 3 | 25 | 34 | 85 |
| $R$ | 26183 | 40004 | 18032 | 5115 |
| $F$ | 0 | 19 | 162 | 609 |

**Table S3:** Fixed parameters.

| Variable | Value | Definition | Reference |
| --- | --- | --- | --- |
| $\theta$ | 7 days | Duration of infectious period | Cevik et al. Lancet Microbe 2021 |
| $\gamma$ | $1/3 \text{ day}^{-1}$ | Inverse of first part of estimated incubation period, the rate of transition from exposed to next stage of infection (either presymptomatic or asymptomatic) | McAloon et al. BMJ 2020, Lauer et al. Annals Int Med 2020 |
| $\zeta$ | $1/2 \text{ day}^{-1}$ | Inverse of second part of incubation period progression from pre-symptomatic to symptomatic infection | Qin et al. Science Adv |
| $\pi$ | 0.8 | Proportion of infections that are symptomatic infection. | Buitrago-Garcia et al. PLOS Medicine 2020 |
| $\eta_I$ | $1/6 \text{ day}^{-1}$ | Hospitalization rate from symptomatic state, from median days to hospitalization | CDC |
| $\rho_H$ | $1/14 \text{ day}^{-1}$ | Recovery rate from hospitalized | CDC |
| $\delta_I$ | $1/24 \text{ day}^{-1}$ | Fatality rate from symptomatic, inverse of median days from symptom onset to death | CDC |
| $\delta_H$ | $1/20 \text{ day}^{-1}$ | Fatality rate from hospitalized, this assumes no overwhelming of healthcare system ICU capacity | CDC |

**Table S4:**
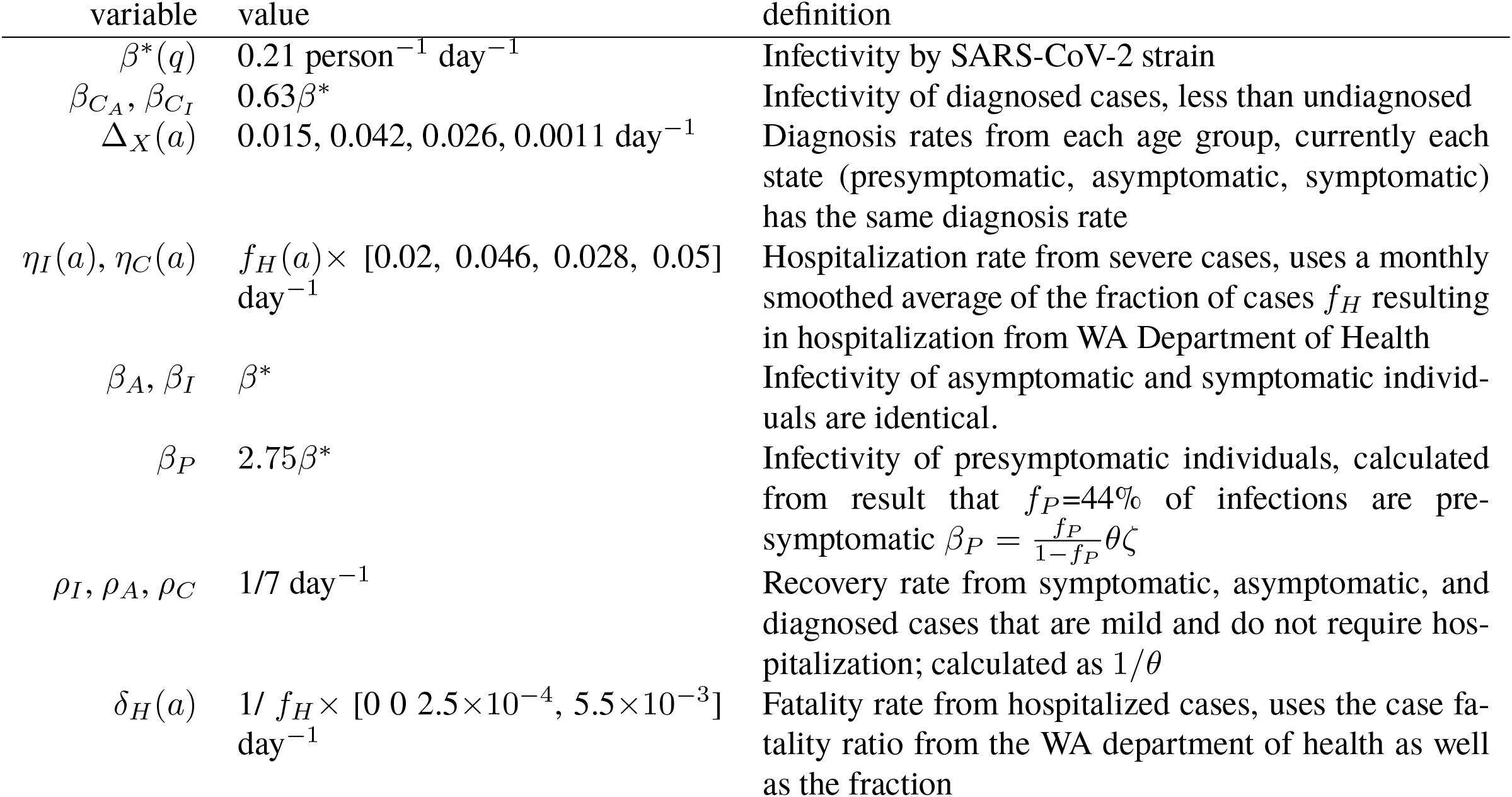
Estimated and inferred parameters.

## Footnotes

1 Buitrago-Garcia *et al*. PLOS Medicine (2020) doi:10.1371/journal.pmed.1003346.

2 Swan, D. A. et al. Vaccines that prevent SARS-CoV-2 transmission may prevent or dampen a spring wave of COVID-19 cases and deaths in 2021. medRxiv 133, 323–57 (2020)

